# Student perceptions of cardiovascular physiology module format in a preclinical medical science course

**DOI:** 10.1101/2023.05.28.23290652

**Authors:** Kristen Scherrer, Spencer Sullivan, Gary Beck Dallaghan, Emily Moorefield

**Author notes:** Corresponding author (EM). These authors contributed equally to this work (Kristen Scherrer, Spencer Sullivan).

## Abstract

Positive active learning outcomes require student engagement with foundational, preparatory material prior to class. The current study involved thorough updating of cardiovascular physiology module format. It then examined preclinical medical student perceptions, and midterm exam performance, after using different preparatory module formats that were reviewed prior to participating in interactive classroom sessions. Modules that were initially created in Articulate Storyline were updated in Articulate Rise360 and introduced over a 3-year period. Both module styles contained the same cardiovascular physiology content, but updated Rise360 modules presented content in multiple formats to capture a variety of student learning preferences and divided each concept into several smaller topics to maintain student attention. Although midterm exam performance remained unchanged, student evaluations revealed that the updated Rise360 modules were more helpful with fewer technical issues indicating that students preferred the interactive online modules to prepare for collaborative classroom exercises. Students find updated preparatory modules to be more helpful and may therefore be more likely to engage with them before class and ultimately lead to a more productive interactive classroom learning experience.

## Introduction

Implementation of active-learning techniques has consistently shown to enhance concept retention and student achievement [1,2]. One approach to active learning, the flipped classroom, requires that students acquire foundational knowledge outside of class and then apply this knowledge through higher order thinking and problem-solving exercises with the support of faculty and peers in class [3, 4]. Thus, student preparation outside of class is essential for successful implementation of this type of active learning technique [5]. Faculty-created modules are one way to provide students with foundational, preparatory content and have the advantage of being customized to cover specific course objectives useful for interactive classroom exercises. Students are responsible for a thorough and thoughtful review of module content prior to class so that they are prepared to actively engage with faculty and peers during the classroom session.

The current study therefore sought to examine student perceptions of preparatory module format when learning basic cardiovascular physiology. An initial set of modules on six cardiovascular physiology concepts was created in roughly the year 2000 using Articulate Storyline, which included audio, video, and matching questions. Module topics included: membrane potential, cardiac cycle, electrocardiogram, vascular hemodynamics, cardiac output, and regulation of mean arterial pressure. Each of the six Storyline module videos had a run time of approximately one hour. Storyline modules were initially well received; however, as time has progressed they have become technologically outdated as they are not compatible with popular internet browsers and do not allow for variable speed playback.

Newer modules were created in Articulate Rise360 on the same six cardiovascular physiology concepts. Although content remained the same, modernized modules presented content in multiple ways including brief videos allowing variable speed playback, corresponding web-based text and graphics, interactive images, and multiple-choice check-your-understanding questions. Updated modules also divided each concept into several smaller topics. For example, cardiac cycle module topics include events of the cardiac cycle, volumes and pressures of the cardiac cycle, heart sounds, and pressure volume loops. Updated modules were introduced in a stepwise manner beginning in 2019, when students were assigned two new Rise360 modules on traditionally more complex topics, the cardiac cycle and cardiac output, and the original Storyline modules on the remaining topics. Then, beginning in 2020, and in 2021, students were assigned updated Rise360 modules on all six basic cardiovascular physiology topics. The 2017-18 school years were included as a comparison group since all five modules assigned were Storyline modules. Herein we examined student, end-of-block evaluations and discovered that students found the updated Rise360 modules to be more helpful and have fewer technical issues than the original Storyline modules.

## Materials and Methods

Student perceptions of module format were assessed by examining end-of-block evaluations. In the evaluations, students rated modules, as well as other modes of instruction, using a 5-point Likert scale ranging from “not at all helpful” to “extremely helpful”, or by selecting, “Did not use/NA to this block”. Student ratings were compared across years using the Kruskal-Wallis Test with Mann-Whitney U test for nonparametric data (IBM SPSS, v24).

In addition, narrative comments that included the word ‘module’ were extracted from open-ended evaluation questions including, “Please comment about your experiences with content and organization.”, and “Which methods and/or materials were most helpful? What could have been more helpful? How?”. Narrative comments were inductively analyzed, and four common themes were identified: organizational, technical, content, and general, which were further categorized as a ‘pro’ comment or a ‘con’ comment (see Figure 1 caption for details). Three independent researchers then deductively categorized themes from the student comments and discrepancies were discussed until full agreement was reached. Data was anonymously collected and analyzed in aggregate.

**Fig 1.**
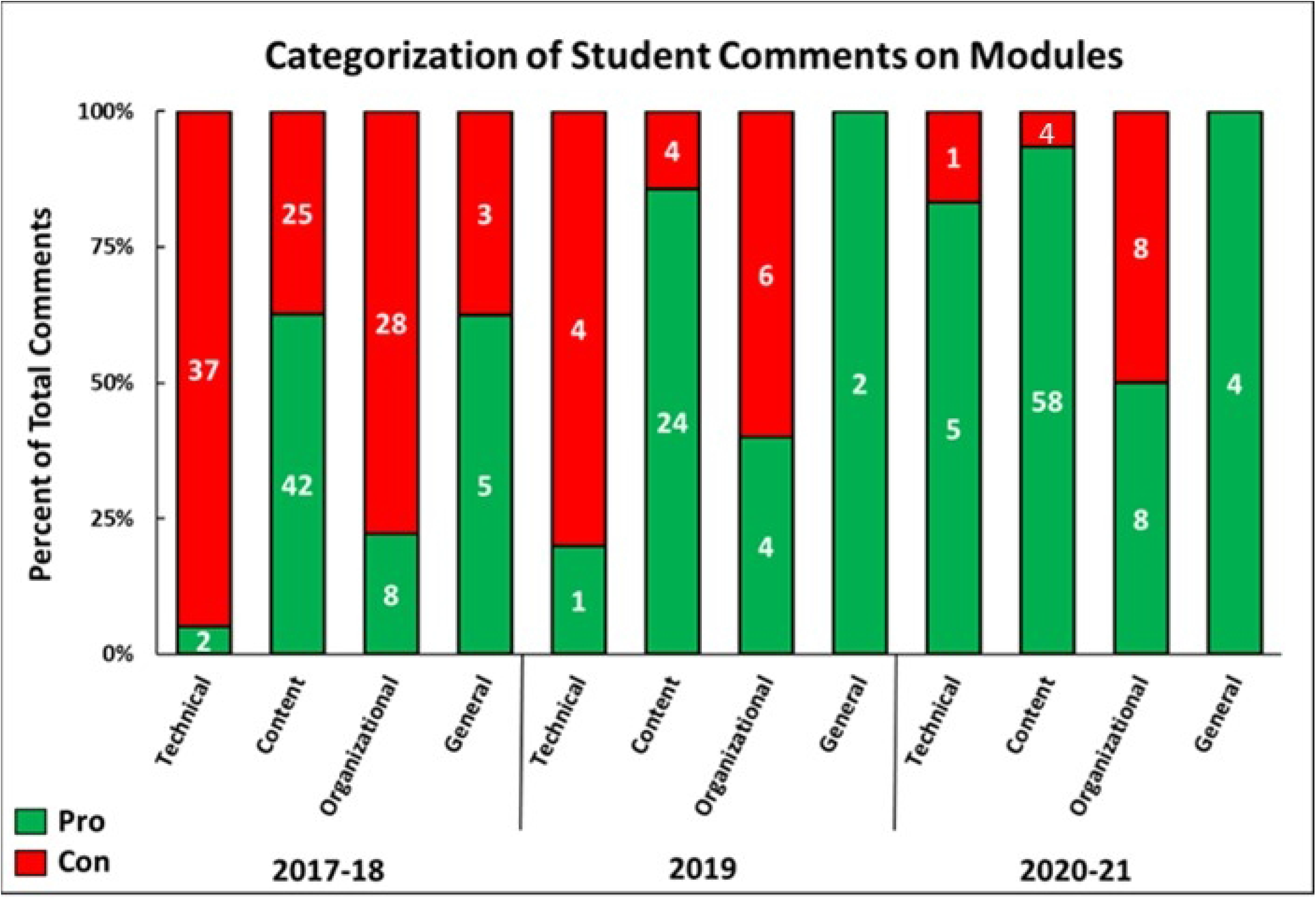
Categorization of open-ended responses containing the word ‘module’ for each student cohort. Data are presented as the percentage of comments in each category (y-axis) as well as the number of comments in each category (numerical values within bars). Comments containing the word ‘module’ were extracted and inductively categorized into four themes: technical, content, organizational, and general. Technical comments discussed technical features like functionality, speed, and browser compatibility. Content comments discussed the usefulness of the modules and other non-technical aspects including the amount of detail in the module and organization of an individual module.

Organizational comments discussed the flow of content and instruction throughout the block including the timing of modules and their connections with other material. General comments discussed non-specific, generalized descriptions of modules. Comments were further divided into ‘pro’ and ‘con’ categories based on whether the overall impression was favorable or not. Some comments included more than one sentiment and were therefore classified into multiple categories.

Additionally, student performance on the cardiovascular block multiple-choice midterm exam was analyzed across each year using the Kruskal-Wallis Test to assess comprehension and retention differences in module format types. The general content and number of questions per topic remained similar from year to year.

This study was approved by the UNC-Chapel Hill Office of Human Research Studies and conducted under the guidelines of UNC IRB Study 22-2111. Consent was not obtained because all data were analyzed anonymously.

## Results

A Kruskal-Wallis test demonstrated that student ratings of module helpfulness increased as additional Rise360 modules were introduced, H(2) = 35.14, p < 0.001 (see Table 1 for descriptive statistics and student ratings by year). Mann-Whitney tests were used to compare all groups and showed that module helpfulness ratings in 2019 were significantly greater compared to 2017-18 (p = 0.022), and in 2020-2021 they were significantly greater compared to 2019 (p = 0.006) and 2017-18 (p < 0.001). These findings suggest students were more satisfied with the quality of updated Rise360 modules compared to original Storyline modules.

**Table 1.** Modules were rated on their level of helpfulness using a 5-point Likert scale by each student cohort, 2017-18, 2019, and 2020-21. * Statistical Significance using Kruskal-Wallis and Mann-Whitney tests (*p* < 0.05). Abbreviations: n, sample size; M, mean; SD, standard deviation.

| Course Year | 2017 - 2018 | 2019 | 2020 - 2021 |
| --- | --- | --- | --- |
| Students per group (n) | 369 | 180 | 356 |
| Module Rating; M (SD) | 3.35 (1.24)* | 3.62 (1.10)* | 3.89 (1.09)* |

Analysis of student comments containing the word ‘module’ revealed an increased number of positive comments in all four categories with the implementation of updated Rise360 modules (Figure 1).

Initially, in 2017-18 when students were assigned only Storyline modules, there was a high number of comments associated with technology and the majority were negative (39 technical comments total; 37 con). These comments referenced the inability to access the modules in Chrome and the inability to view the module at 2x speed, among other functional issues. Over the 3-year incorporation of the Rise360 modules, the total number of technical comments decreased dramatically (37 con comments in 2017-18 down to 1 con comment in 2020-21) with mostly positive comments by 2020-21 (6 technical comments total; 5 technical pro) and reference to the ease of use.

The overall number of comments regarding module content did not change drastically during the module updating period. However, interestingly, the percentage of positive comments for module content increased each year and in 2020-21, when all the modules assigned were Rise360 format, 85% of module content comments (75 of 88 total) were positive even though the content presented remained the same. Student comments regarding organization of content within the block including the timing of modules and their relationship to other material also decreased in number over time and there were many fewer negative organizational comments with the introduction of Rise360 modules. General comments about the modules did not reference specific features and showed a similar trend with fewer general con comments after incorporating the Rise360 modules.

Analysis of midterm exam performance across years failed to show any significant difference: 81.2% (SD 8.4), 78.9% (SD 10.8), 80.7% (SD 9.5) for 2017-18, 2019, and 2020-21, respectively, (H(2) = 4.75, p = 0.093). This finding demonstrates that the newer modules did not directly influence immediate student learning or retention.

## Discussion

In this study cardiovascular physiology modules were thoughtfully updated with the goal of increasing student engagement and understanding of foundational cardiovascular physiology concepts. Original Storyline modules presented each of six cardiovascular physiology concepts as a single video with a run time of approximately one hour, a format which may be challenging or overwhelming for students to process and comprehend. Updated Rise360 modules presented topics using multiple learning modalities, which appeals to a variety of learning preferences and encourages a wide range of students to interact with content. In addition, Rise360 modules separated each concept into smaller parts to allow easier navigation, prevent distraction, and facilitate completion without interruption.

Our analysis of student evaluation ratings and comments revealed that Rise360 modules were more helpful and had fewer technical issues than the original Storyline modules. Previous research demonstrates that interactive modules increase student study time [6] which may enhance active learning classroom activities. In addition to study time, increased usage may also translate into a greater number of students that engage with the modules at a time when engagement is dwindling [7]. One limitation of the current study; however, is that our institution’s learning management system was not configured to track the number of students that opened modules or the amount of time each student spent interacting with the modules. Although student evaluations suggest there was more engagement with the updated modules, we are unable to directly compare student interaction between the different module styles.

Interactive modules that provide learner control, graphics and videos, and self-assessments have shown to significantly increase medical student learning of basic science concepts [8, 9]. The present study confirms that students found the updated Rise360 modules containing these features to be more helpful. However, regardless of the module format assigned, student performance on the cardiovascular midterm exam remained unchanged. Our findings are consistent with other mixed results on the impact of modules [6] and flipped classrooms [4] on promoting knowledge acquisition and retention.

One complicating factor in our study is that overall exam performance has declined over the past several years, perhaps due in part to the stress of the COVID-19 pandemic. Pandemic-related stress has shown to negatively impact mental health and exam performance in medical students [10, 11]. It is possible to consider that students’ cardiovascular midterm exam scores may have also significantly declined if not for the updated Rise360 modules. Additional research is needed to determine whether this may be the case and to further investigate specific module features that may enhance student learning.

In conclusion, our findings suggest that careful consideration of preparatory material format is critical for maximum student satisfaction in preparation for active learning classroom activities. Presenting preparatory modules in a modern, interactive format with self-assessment improves student perceptions of the materials. Students find updated preparatory modules to be more helpful and may therefore be more likely to engage with them before class, which may lead to a more productive interactive classroom learning experience.

## Data Availability

Data files are available from the figshare database (https://figshare.com/s/3d90323da8c6710b7ac8).

